# Bridging the gap: A qualitative analysis of online public forum with healthcare professionals in bursting myths about weight management

**DOI:** 10.1101/2022.06.03.22274963

**Authors:** R Bhatti, M Joumaa, B Bereczky

**Affiliations:** Consultant Endocrinologist, Mediclinic City Hospital, Dubai, UAE; Dietician and bariatric co-ordinator, Mediclinic Parkview Hospital, Dubai, UAE; Consultant bariatric surgeon, Mediclinic Dubai Mall, Mediclinic Parkview and Mediclinic City Hospital, Dubai, UAE

**Keywords:** obesity, zoom meeting, weight management

## Abstract

**Background:** Obesity has increased rapidly worldwide in last few decades. According to World Obesity Federation 2018, the prevalence of obesity in UAE amongst Emirati aged 18-69 years has been reported as 32.2% vs 38.5% living with obesity vs overweight in men and 41.8% vs 29.3% living with obesity vs overweight in women. There are many myths amongst general public about obesity.

**Aim:** The objective of the study was to address public questions through online forum regarding weight management.

**Methods:** In this cross-sectional observational study qualitative analysis was done. It was conducted quarterly at Mediclinic City and Parkview Hospital in Dubai from August 2020-September 2021. An online zoom meeting was made available to the public. People who wanted to attend the session registered via dedicated email address. A certified dietician, endocrinologist and bariatric surgeon were available to answer people’s questions.

**Results:** 6 online forums have been conducted. 163 (range 10-90) people attended. Three themes were constructed during data analysis. 1-Diet: what’s the best diet and about caloric intake. 2-Insulin resistance or hormone imbalance is responsible for weight gain. 3-Misinformation and unrealistic expectations about bariatric surgery. Feedback from audience was ‘it was interactive, fun and informative session’, ‘it’s good to know that it’s not my fault and I need to get help’.

**Conclusion:** Our results show that patients need support throughout their weight management journey. Healthcare professionals need to be more aware of patient’s perceptions about weight management so that they can be addressed in consultations. We need more online support sessions facilitated by healthcare professionals.

## Introduction

The prevalence of obesity has reached pandemic levels worldwide. According to World Obesity Federation 2018, the prevalence of obesity in UAE amongst Emirati aged 18-69 years has been reported as 32.2% vs 38.5% living with obesity vs overweight in men and 41.8% vs 29.3% living with obesity vs overweight in women. (1) The prevalence of people living with overweight and obesity, by BMI, were reported as 43.0 and 32.3%, respectively in the UAE. (2)

The Awareness, Care, and Treatment in Obesity maNagement-International Observation ACTION-IO study investigated the perceptions, attitudes, and behaviors of people with obesity and healthcare professionals. (3) In UAE 76% of people with obesity admitted to full responsibility for their weight loss; 84% of healthcare professionals acknowledged responsibility for actively contributing to patient weight loss efforts. Both people with obesity and healthcare professionals acknowledged obesity as a chronic disease however, several misperceptions were identified that may be limiting current obesity management in the UAE.

COVID 19 pandemic with consequent restrictions on staying or working from home has changed how people were communicating with each other. This has resulted in increased use of video conferencing as a means of communicating e.g Zoom meetings. (4,5) The aim of this study was to address public questions through online Zoom forum regarding weight management.

## Methods

### Study Design

It was a cross-sectional observational study. It was conducted quarterly at Mediclinic City and Parkview Hospital in Dubai from August 2020-September 2021. An online zoom meeting was made available to the public. The meeting was held for 1 hr. The link was sent to people who were registered at Mediclinic Hospital database. People who wanted to attend the session registered via dedicated email address. A certified dietician, endocrinologist and bariatric surgeon were available to answer people’s questions.

### Data collection

All interviews were audio- and video recorded, and participants reaffirmed consent verbally prior to Zoom recording. All questions received either in email prior to meeting, asked during zoom session or entered in comments box during the zoom meeting were included in qualitative analysis. All three healthcare professionals individually looked at the questions.

### Ethics Statement

Ethics approvals were obtained from the local Mediclinic Institutional Research Board and the Dubai Scientific Research Ethics Committee, Dubai Health Authority.

## Results

6 online public Zoom forums have been conducted during the study period. 163 (range 10-90) people attended. Graph 1 shows the attendance of participants in each session.

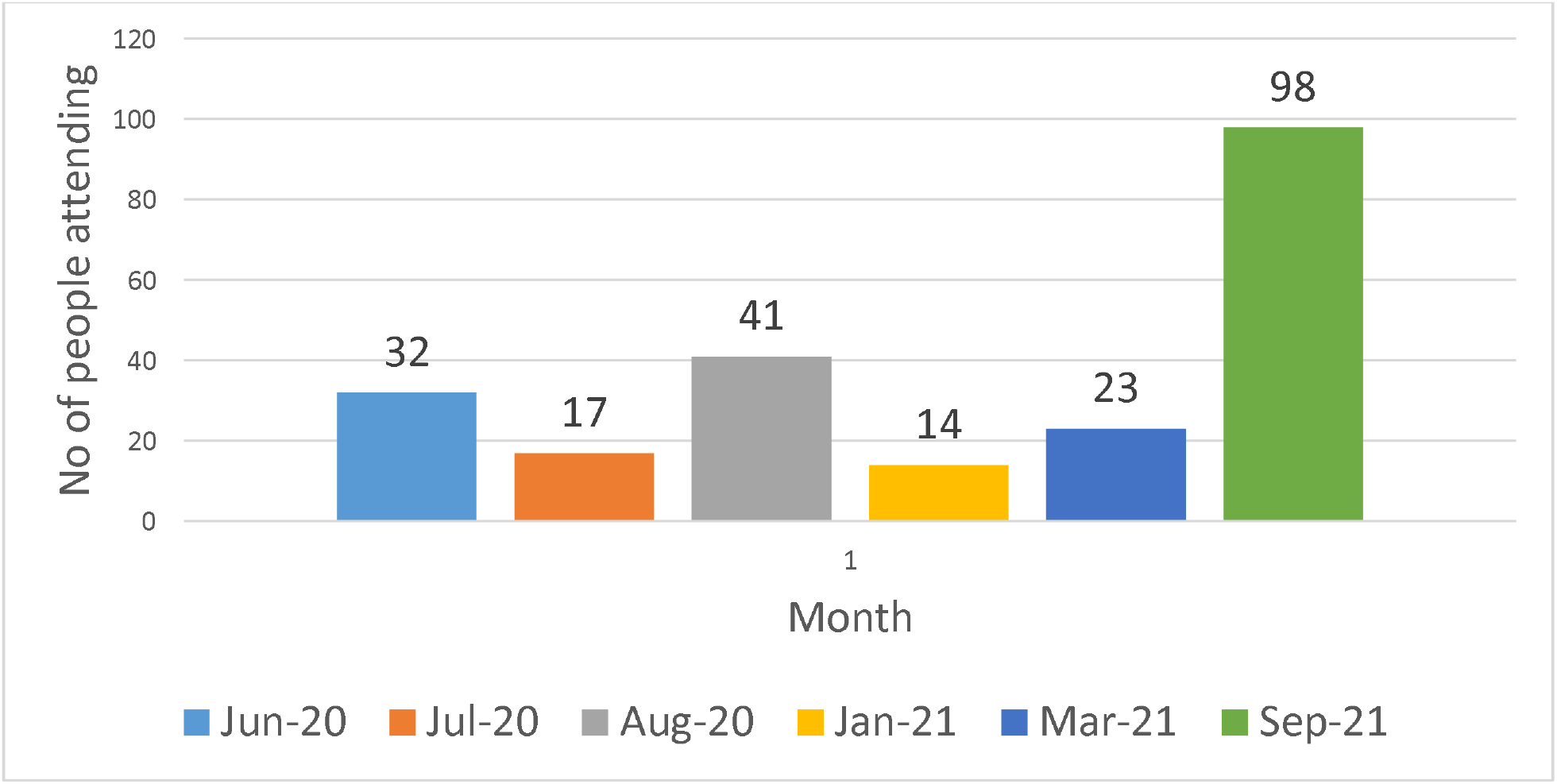

### Three themes were constructed during data analysis

#### Diet

People were really interested about how diet can help to lose weight and how it works?

“what’s the best diet and about caloric intake”

“What is the best diet and exercise for insulin resistant individuals?” “What would be the reason behind the weight plateau?”

“At what time should be the last meal is taken?” “Can you share a good diet plan for weight loss?”

“How to lose weight without the feeling of restriction”

“What to do about cravings for sugar?”

What are the vitamins that should be taken during dieting and exercising to lose weight?”

“Foods to replace oestrogen in menopause”

### Insulin Resistance /Hormone Imbalance

People had this perception that weight gain and their difficulty in losing weight was linked to thyroid problem or insulin resistance.

- “How can you lose weight with Hashimoto’s disease?”
- “What food is best to speed up metabolism and to help hypothyroidism?”
- “I have obesity issues due to chronic PCOS”
- “Why it hard to lose weight when you are at your late 40s despite eating healthy and regular exercise”
- “Can you do intensive work out if you suffered with hypertension and asthma?”
- “How to avoid weight gain during menopause?”

### Misinformation and unrealistic expectations about bariatric surgery

Some people hadn’t even heard about bariatric surgery or when they should be looking at it as one of the options in their weight management journey.

- “How to develop muscles after bariatric surgery?”
- “My BMI is 30 and I am already doing exercise and diet can I go for surgery?”
- “How much weight can I lose after bariatric surgery?”
- “How to target my belly fat?”
- “What is bariatric surgery?”

The three healthcare professionals answered people’s questions regarding their weight management. Patients were shown few slides on Microsoft PowerPoint to explain some of the concepts of obesity. People were also guided who to contact and get further assessment by their healthcare professionals.

### Feedback from People

The feedback about these sessions from patients was really positive.

- ‘It was interactive, fun and informative session’.
- ‘It’s good to know that it’s not my fault and I need to get help’.
- “Thank you very much, this was really informative”.

## Discussion

Our study has identified three different themes from these sessions. People have different perceptions about diet, how it works and looking for best meal plans to help them lose weight. It also did show about people’s frustration about weight regain once they go back on normal diet. These findings are similar to myths and perceptions discussed by Krista et al. (6) The certified dietician addressed these questions on diet and explaining to patients the concepts of energy intake and output, solving the weight plateau etc. This was done according to patient centered guidelines for adult obesity management. (7)

The second theme was regarding people’s perception that it’s metabolic problem or hormonal imbalance which is contributing to their weight gain. This perception of link of hypothyroidism and obesity has been well documented before. (8)

The third theme was perceptions and misconceptions about bariatric surgery. Some people were not even aware of what bariatric surgery is? Some thought that it was last resort in weight management. One patient quoted

“Bariatric surgery is extreme measure, it’s scary and have I reached that stage’ Mikaela et al had described similar results in analysis of their online forum discussions about bariatric surgery. (9)

There is still insufficient literature including randomized controlled trials on use of social media for use of online support for patients and healthcare professionals. A scoping review of 284 studies including 48 randomized controlled trials 35.4% reported statistically significant results favoring the social media intervention being evaluated. It included studies on lifestyle/weight loss and cancer. (10) However, the advantage of zoom session is that it can be anonymous communication as person doesn’t have to switch on video but can still ask question in comment box. It helps in reducing stigmatization associated with obesity. (11) The concept of using zoom public meetings is not new as Katherine et al reported that participants in online forums are quite similar to those in in-person ones. (12)

This study has highlighted the perceptions that people have in terms of their weight, diet, hormones and bariatric surgery. Healthcare professionals need to play an important role in addressing these perceptions and guiding people in right direction. As it’s cross sectional study it’s difficult to do in-depth analysis into answers. Also there are no follow up appointments from these zoom sessions to address people’s questions in depth. More studies using this medium of communication are needed to assess their utilization in clinical management of obesity.

## Conclusion

- Our results show that patients need support throughout the weight management journey.
- Healthcare professionals need to be more aware of patient’s perceptions about weight management so that they can be addressed in consultations.
- We need more online support sessions facilitated by healthcare professionals.

## Data Availability

All data in the work is in the manuscript

